# LOMIA-T: A Transformer-based LOngitudinal Medical Image Analysis framework for predicting treatment response of esophageal cancer

**DOI:** 10.1101/2024.03.29.24305018

**Authors:** Yuchen Sun, Kunwei Li, Duanduan Chen, Yi Hu, Shuaitong Zhang

## Abstract

Deep learning models based on medical images have made significant strides in predicting treatment outcomes. However, previous methods have primarily concentrated on single time-point images, neglecting the temporal dynamics and changes inherent in longitudinal medical images. Thus, we propose a Transformer-based longitudinal image analysis framework (LOMIA-T) to contrast and fuse latent representations from pre- and post-treatment medical images for predicting treatment response. Specifically, we first design a treatment response-based contrastive loss to enhance latent representation by discerning evolutionary processes across various disease stages. Then, we integrate latent representations from pre- and post-treatment CT images using a cross-attention mechanism. Considering the redundancy in the dual-branch output features induced by the cross-attention mechanism, we propose a clinically interpretable feature fusion strategy to predict treatment response. Experimentally, the proposed framework outperforms several state-of-the-art longitudinal image analysis methods on an in-house Esophageal Squamous Cell Carcinoma (ESCC) dataset, encompassing 170 pre- and post-treatment contrast-enhanced CT image pairs from ESCC patients underwent neoadjuvant chemoradiotherapy. Ablation experiments validate the efficacy of the proposed treatment response-based contrastive loss and feature fusion strategy. The codes will be made available at https://github.com/syc19074115/LOMIA-T.

## 1 Introduction

The analysis of longitudinal medical images is an important but challenging problem in the monitoring of tumor progression and the evaluation of treatment response [1, 2]. Compared to single time-point medical images, longitudinal medical images offer more information related to treatment response [3]. For instance, neoadjuvant chemoradiotherapy (nCRT) followed by surgical resection is the preferred treatment for locally advanced esophageal squamous cell carcinoma (ESCC) [4]. For patients who achieved pathological complete response (pCR) after nCRT, a wait-and-see strategy is more suitable than surgical resection [5]. Therefore, preoperative prediction of pCR for individual ESCC patients is highly desirable, as it can assist clinicians in making treatment decisions.

Previous studies have demonstrated using deep learning-based or radiomics-based methods to analyze contrast-enhanced CT images can preoperatively predict pCR of ESCC [6]. Nevertheless, the majority of studies primarily concentrate on single time-point CT images (pre- or post-treatment), neglecting the temporal dynamics and alterations which can be elucidated through longitudinal contrast-enhanced CT images [7]. Several methods for predicting treatment response based on longitudinal medical images have been proposed, demonstrating that disease progression patterns represented by longitudinal data can improve the performance [8, 9]. These works can be primarily categorized into deep feature contrast (DFC) based methods and deep feature fusion (DFF) based ones. Both of them usually establish two or more parallel neural networks to represent longitudinal images. Afterward, the DFC-based methods utilize a similarity function to quantify the differences among features from longitudinal images [10, 11], which are usually correlated to the treatment response. It is similar to the way clinicians evaluate tumor treatment response where they typically rely on visual comparison between pre- and post-treatment CT images. Thus, DFC-based methods are clinically interpretable, which is crucial for clinical decision-making. In contrast, the DFF-based methods usually fuse the features from longitudinal images and then associate the fused feature with treatment response. Compared with DFC-based methods, DFF-based methods for longitudinal images can provide a more comprehensive and accurate analysis of longitudinal changes [12, 13]. Common feature fusion strategies include concatenation of features with all- or cross-attention mechanism [14], and their effectiveness have been validated in various tasks. For example, Tong et al. [15] developed a Dual-input Vision Transformer (DiT) model with the all-attention and concatenation strategy to fuse features from pre- and post-treatment medical images in breast cancer patients. However, self-attention leads to each output token encompassing features from every token in the longitudinal images. Concatenating all longitudinal tokens directly may introduce redundancy into the final feature pool, posing a potential impact on predictive performance. Disentangled representation learning is a method reducing the redundancy in fused feature pool with aligning longitudinal data in the time dimension [16] or latent representation dimension [17] to capture meaningful aspects. For instance, Yue et al. [18] proposed Multi-loss disentangled representation learning (MLDRL) to highlight differences and align commonalities among longitudinal images.

In this paper, we introduce LOMIA-T, a novel longitudinal image analysis framework that combines feature contrast and fusion techniques, through which, we can effectively leverage longitudinal medical images for predicting treatment response. Specifically, we develop a clinically interpretable fusion strategy for longitudinal medical images and introduce a treatment response-based contrastive loss function aimed at capturing nuances in disease progression trajectories. Experimental results on an in-house ESCC dataset from two hospitals demonstrate that LOMIA-T outperforms other state-of-the-art longitudinal image analysis methods. Furthermore, LOMIA-T can be transferred for predicting treatment response in other diseases and can accommodate multiple time-point medical images with minor modifications.

## 2 Materials and Methods

### 2.1 CT Imaging and Preprocessing

This study was approved by the ethics committee of the participated hospitals and conducted in accordance with the ethical standards of the Helsinki Declaration. A total of 170 locally advanced ESCC patients who underwent nCRT are obtained from two hospitals, and longitudinal contrast-enhanced CT images (pre- and post-treatment) for these patients are available. We randomly divide these patients into a training and a test set at a ration of 4 : 1. We first use nnU-Net [19] to segment tumor region on the pre- and post-treatment contrast-enhanced CT images automatically (Figure 1A). To guarantee the acquisition of the whole tumor region, we expand the three-dimensional bounding box of the segmented mask by 4 pixels on the cross-sectional slices and select 32 consecutive slices as the region of interest (ROI).

**Fig. 1.**
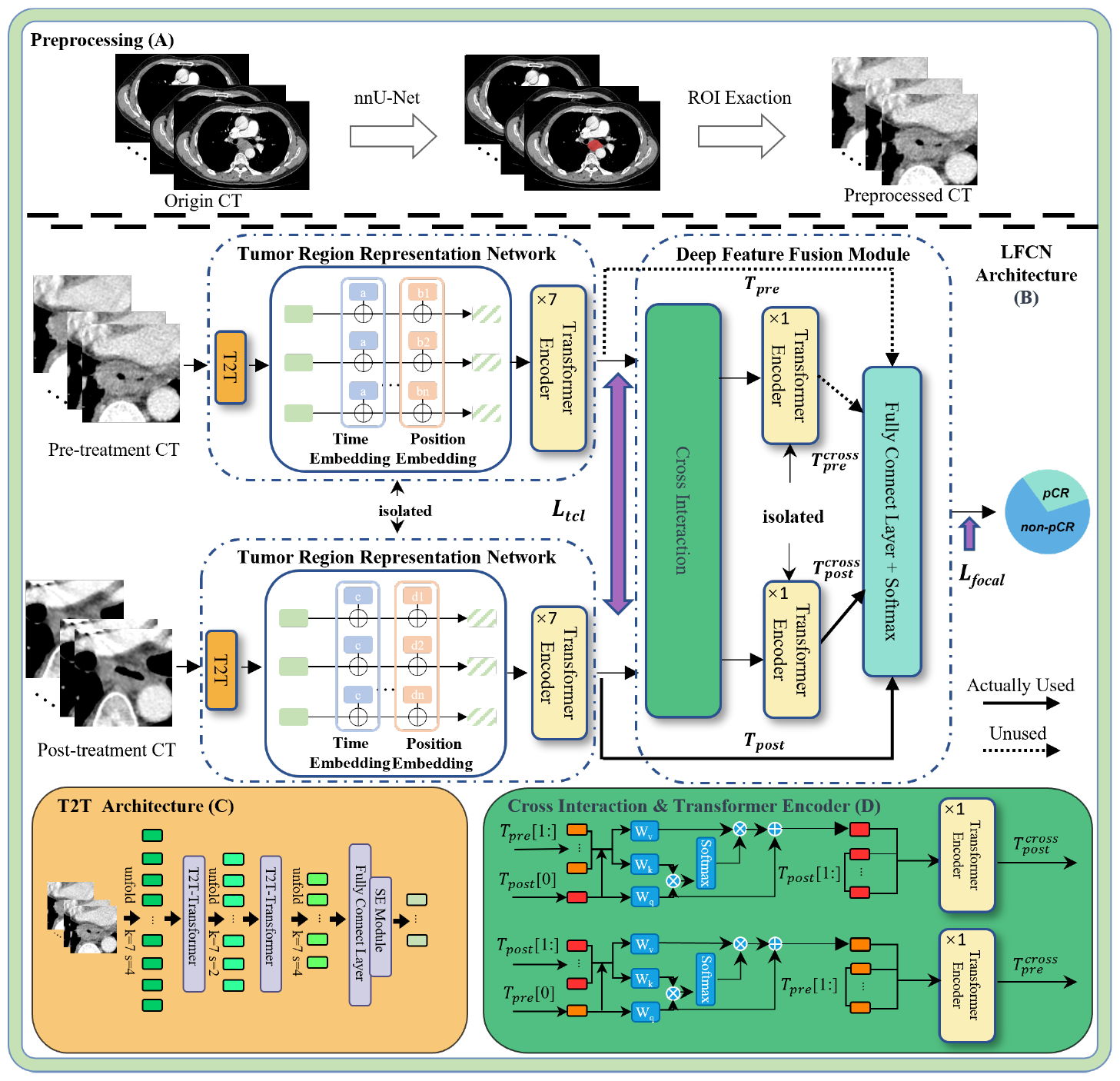
Schematic of LOMIA-T. Solid lines represent *T*_*post*_ and 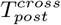 which are used in LOMIA-T, while dashed lines represent *T*_*pre*_ and 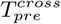 which are not.

### 2.2 Overall Network Architecture

Fig. 1B illustrates the network architecture of LOMIA-T. It mainly contains two subnetworks: tumor region representation network and deep features interaction network for pre- and post-treatment CT images. We propose a treatment response-based contrastive loss, enhancing the ability of the tumor region representation network to discern feature disparities indicative of treatment effects between pre- and post-treatment CT scans. Furthermore, a novel deep feature fusion method is introduced to improve prediction performance further.

#### Tumor region representation network

The tumor region representation network is constructed using a token-to-token Vision Transformer (T2T-ViT) module, and the representation networks for pre- and post-treatment contrast-enhanced CT images are structurally identical and mutually independent. T2T module is utilized to transform the input *X*_*pre*_, *X*_*post*_ ∈ ℝ^*D*×*W* ×*H*^ (D = 32, W = 48, and H = 48 here) into tokens instead of the hard split used in the ViT [20] because it can retain textural and structural information. Each T2T module involves three steps: soft split (SS), T2T-Transformer (TT), and re-structurization (RS), with the step size in the soft split set to (4, 2, 2) and the same scale as the ROI while re-structurization. The output dimension in the T2T module is halved using a fully connected layer to reduce the complexity of the representation network and improve its generalization ability. Figure 1C illustrates the T2T structure. The iterative process in T2T module can be formulated as:

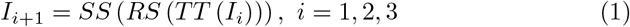

We apply a soft split at first to split *X*_*pre*_, *X*_*post*_ to tokens: *I*_1_ = *SS* (*X*_*i*_), *i* = *pre, post*. The final iteration results in the output tokens 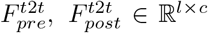 where *c* represents the number of channels and *l* signifies the output dimension.

In longitudinal images modelling, spatial and feature misalignments of the ROI on the longitudinal images always occur [21]. To mitigate spatial misalignment, two learnable matrices *PE*_*pre*_, *PE*_*post*_ ∈ ℝ^*l*×*c*^ are introduced as positional embedding, respectively. Positional embeddings describe the spatial relationships in images, while temporal embeddings indicate the temporal changes in images. Previous methods involve temporal embedding usually introduce a different temporal variable for different tokens [15]. We assume that tokens from the same time-point image share the same temporal embedding. Here, two learnable temporal embedding variables *TE*_*pre*_, *TE*_*post*_ ∈ ℝ^1^ are also introduced for 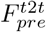 and 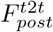, respectively. After spatial and temporal embeddings, tokens from pre- and post-treatment CT images can be expressed as:

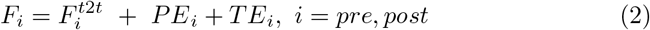

Where *F*_*pre*_, *F*_*post*_ ∈ ℝ^*l*×*c*^.

We map *TE*_*pre*_, *TE*_*post*_ ∈ ℝ^1^ to ℝ^1×*c*^ using the Python-specific broadcasting operation and then add them together. Afterward, a seven-layer transformer encoder is employed to extract high-level imaging features *T*_*pre*_, *T*_*post*_ ∈ ℝ^*l*×*c*^.

#### Treatment response-based contrastive loss (TCL)

Contrastive loss aims to reduce the distance between similar input pairs and increase that between dissimilar pairs in the feature space [22, 23]. For all ESCC patients, CT imaging features will change after nCRT. However, pCR group (with no tumor cells remained after nCRT) experiences a totally tumor regression, while non-pCR group experiences a smaller tumor regression or even tumor progression. In the task of predicting pCR of ESCC patients, the pre- and post-treatment CT images for patients who achieve pCR are considered as dissimilar pairs, and the pre- and post-treatment CT images for patients who does not achieve pCR are considered as similar pairs. Accordingly, we design a treatment respones-based contrastive loss (TCL) for predicting pCR:

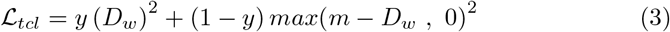

Where *y* represents the patient’s treatment response label (pCR = 0, non-pCR = 1), *D*_*w*_ =∥ *T*_*pre*_ *- T*_*post*_ ∥_2_ denotes the Euclidean distance between features from pre- and post-treatment CT images. For pCR patients, we used the penalty only if *D*_*w*_ is smaller than *m* where *m* equals 0.5.

#### Deep Feature Fuison Module

The fusion module integrates tumor representations from pre- and post-treatment CT images using a cross-attention mechanism [14], as illustrated in Fig. 1D.

Specifically, for post-treatment CT branch, it first collects the patch tokens (*T*_*pre*_[1 :]) from the pre-treatment CT branch and concatenates its own CLS tokens (*T*_*post*_[0]) to *T* ^*′post*^ ∈ ℝ^*l*×*c*^. The mechanism then performs cross-attention between 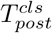 and *T* ^*′post*^ with multiple heads (MCA), where CLS token is the only query as the information of patch tokens are fused into CLS token. The output 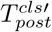 with layer normalization and residual shortcut is defined as follows:

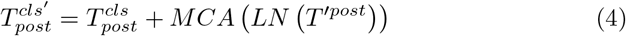

Afterward, the CLS token and patch tokens from post-treatment CT branch are fed into a single layer transformer encoder including multi-head self-attention (MSA) mechanism and feed-forward network (FFN):

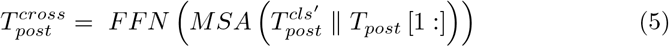

Where 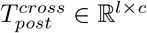.

Considering the redundancy between 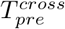 and 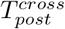, their direct concatenation may be not the optimal feature fusion strategy. Previous study demonstrated that the combination of post-treatment CT imaging features and their interaction features with pre-treatment CT imaging features perform best for predicting pCR [24], which is consistent with the perception of clinicians. Inspired by the clinical prior knowledge and our previous finding, we concatenate 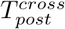 and *T*_*post*_ with skip connection for predicting pCR. In this way, LOMIA-T can better utilize low and high level semantic information for predicting pCR. Specifically, the first token in 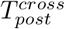 and *T*_*post*_ is considered as the CLS token, respectively. These two CLS tokens are then processed through a fully connected layer with Softmax activation to create a binary classifier, enabling the computation of pCR classification results. In subsequent ablation studies, we also compare the performance of our fusion strategy with others.

Focal loss [25] is used as loss function to emphasize challenging samples here, denoted as *ℒ*_*focal*_:

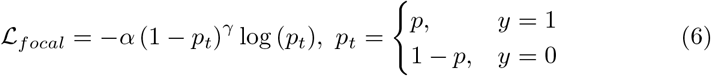

Where, *y* = 1 represents the non-pCR class, *y* = 0 represents the pCR class and *p* ∈ [0, 1] is the probability for the class with label *y* = 1. Here, *α* balances the importance of pCR/non-pCR examples, and *γ* represents a tunable focusing hyperparameter. Additionally, *α* and *γ* are set to [1,1] and 2, respectively. The overall loss *ℒ* is a weighted combination of *ℒ*_*focal*_ and *ℒ*_*ccl*_, and *w*_1_ is set to 0.01 to ensure that these two losses are maintained at a comparable scale.

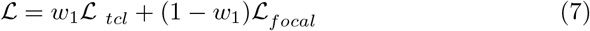

## 3 Results and Discussion

### 3.1 Experimental Settings and Evaluation

We train the LOMIA-T model using ten-fold cross-validation for 100 epochs (5 warm-up epochs) on A100 GPU with a batch size of 64. Other setup includes a cosine linear-rate scheduler with linear warm-up, an initial learning rate of 0.002, a weight decay of 0.05 and a DropKey [26] of 0.4. Moreover, we also apply the individual discrimination task to pre-train the tumor region representation net-work for pre- and post-treatment CT images, respectively [27]. Area under the curve (AUC), accuracy (ACC), sensitivity (SEN), and specicity (SPE) are used to evaluate the classification performance on the test set.

### 3.2 Comparison with Existing Methods

When comparing our proposed model LOMIA-T with three other deep learning methods for longitudinal images (DiT [15] and Siamese-CNN [28], implemented using their official codes, and MLDRL [18], with the experimental results sourced from the original study), LOMIA-T achieves superior performance in predicting pathological complete response (pCR), as illustrated in Table 1. Specifically, LOMIA-T yields an area under the curve (AUC) of 0.886, significantly outperforming DiT and Siamese-CNN (*p* = 0.009 and 0.006, respectively; DeLong test). It is important to note that parameters of DiT and Siamese-CNN are meticulously adjusted to optimize prediction performance using ten-fold cross-validation. Specially, Siamese-CNN adopts the framework of MoCo v2 [28], with a tumor progression task as pretext, which is associated with pCR labels. Additionally, to ensure comparability in model parameters with LOMIA-T, ResNet-18 is utilized as the backbone for Siamese-CNN. We train Siamese-CNN for 100 epochs, with the TCL as loss function. The optimal threshold *m* in the loss function is set to 1.4 using the same approach as that for LOMIA-T.

**Table 1.** Comparison of our model with existing methods for predicting pCR.

| Method | AUC | ACC | SEN | SPE |
| --- | --- | --- | --- | --- |
| LOMIA-T (Ours) | 0.886 | 0.824 | 0.800 | 0.857 |
| DiT [15] | 0.727 | 0.677 | 0.550 | 0.857 |
| MLDRL [18] | 0.866 | 0.810 | 0.684 | 0.875 |
| Siamese-CNN [28] | 0.529 | 0.588 | 0.750 | 0.357 |

### 3.3 Ablation Study

Several ablation experiments are also performed to verify the effectiveness of deep feature fuison strategy and treatment response-based contrastive loss, and the results are listed in Table 2. The training strategies and hyperparameters settings for the ablation studies are the same as LOMIA-T.

**Table 2.** Ablation studies on different strategies for expert classification and effectiveness of TCL loss function on longitudinal ESCC 3DCT dataset.

|  | Strategies | AUC | ACC | SEN | SPE |
| --- | --- | --- | --- | --- | --- |
| (a) | $T_{post} - T_{post}^{cross}$ (LOMIA-T) | 0.886 | 0.824 | 0.800 | 0.857 |
| (b) | $T_{post} - T_{pre}^{cross}$ | 0.879 | 0.765 | 0.750 | 0.786 |
| (c) | $T_{pre} - T_{post}^{cross}$ | 0.850 | 0.765 | 0.750 | 0.786 |
| (d) | $T_{pre} - T_{pre}^{cross}$ | 0.864 | 0.735 | 0.750 | 0.714 |
| (e) | $T_{pre}^{cross} - T_{post}^{cross}$ | 0.843 | 0.735 | 0.650 | 0.857 |
| (f) | w/o Deep Feature Fusion module | 0.711 | 0.735 | 0.850 | 0.571 |
| (g) | Only pre-treatment images | 0.546 | 0.500 | 0.350 | 0.714 |
| (h) | Only post-treatment images | 0.693 | 0.559 | 0.450 | 0.714 |
| (i) | w/o TCL | 0.821 | 0.677 | 0.600 | 0.786 |

#### The effectiveness of deep feature fusion strategy

For predicting pCR, we employ a range of strategies to integrate features from pre- and post-treatment CT images, including the fusion of the features from the pre-/post-treatment after cross interaction 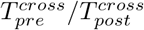 and features from pre-/post-treatment before cross interaction *T*_*pre*_*/T*_*post*_. Our experiments demonstrate that fusing features from *T*_*pre*_ or *T*_*post*_ with those from 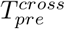 or 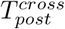 yield superior performance compared to the fusion of 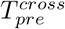 and 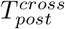, the common strategy employed in previous studies [15]. This may be attributed to the observation that a higher degree of redundant information exists between tokens 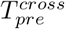 and 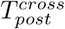 after cross-attention. When comparing the results presented in row (a) and (c), (b) and (d) in Table 2, we observe that imaging features extracted from post-treatment CT images exhibit greater predictive value than those from pre-treatment images. Moreover, in comparison to pCR prediction models based solely on pre- /post-treatment CT images, models integrating both pre- and post-treatment CT images consistently outperform them across all fusion strategies. These findings underscore the importance of longitudinal data in enhancing predictive accuracy for pCR.

#### The effectiveness of treatment response-based contrastive loss

We remove the TCL and employ only focal loss to train the LOMIA-T. Compared with the focal loss alone, the AUC and accuracy values are increased by 6.5% and 14.7%, respectively after adding the TCL. Using TCL forces the tumor representation network to extract pCR-associated imaging features from pre- and post-treatment CT images, thereby improving the pCR prediction performance.

## 4 Conclusion

In this study, we present a Transformer-based framework for longitudinal image analysis to predict treatment response of ESCC patients. We propose a clinically interpretable fusion strategy of longitudinal medical images and experimental results demonstrate its superiority over direct concatenation strategy. Moreover, the proposed treatment response-based contrastive loss can facilitate the model to capture the longitudinal changes of tumor on CT images, leading to a further prediction improvement. The LOMIA-T outperforms other state-of-the-art longitudinal image analysis methods on an in-house ESCC dataset from two hospitals and can be transferred for predicting treatment response in other diseases. In the future, we will further investigate the generalization of LOMIA-T on other diseases and the potential of LOMIA-T in predicting treatment response using multiple time-point medical images.

## Data Availability

All data produced in the present study are available upon reasonable request to the authors

## Acknowledgments

This study is supported by Beijing Natural Science Foundation (L232132) and National Nature Science Foundation of China (82102140).

## Notes

### Competing Interest Statement

The authors have declared no competing interest.

### Author Declarations

This study was approved by the Ethics Committee of the Fifth Affiliated Hospital of Sun Yat-sen University (2021-K62-1) and was conducted in accordance with the Declaration of Helsinki. Informed consent was waived due to the observational design of this study and the de-identifed nature of the data.

